# A Real-world Evaluation of Longitudinal Healthcare Expenses in a Health System Registry of Type-2 Diabetes Mellitus and Cardiovascular Disease Enabled by the 21st Century Cures Act

**DOI:** 10.1101/2025.05.28.25328527

**Authors:** Lovedeep S Dhingra, Aline F Pedroso, Arya Aminorroaya, Jigar Rajpura, Sherif Mehanna, Ivy Tonnu-Mihara, Rohan Khera

## Abstract

**Introduction:** Type 2 diabetes (T2D) is associated with substantial healthcare spending, but quantifying these expenses has been limited to cohorts of self-selected patients or assessments of insurance claims for major healthcare events. Leveraging the 21^st^ Century Cures Act, which mandated reporting hospital-level service, we pursued a comprehensive evaluation of healthcare spending in a diverse cohort of individuals with T2D.

**Methods:** We designed a pragmatic, observational cohort study of patients with T2D seeking regular care (≥ 1 visit/2 years) across 5 hospitals and an outpatient network (2013-2023) in the Yale New Haven Health System. We used the chargemaster file to extract Medicare and insurance-negotiated cash prices for all healthcare events. We used residential zip codes to define median household income based on US Census data. We also examined the prevalence of financial hardship, defined by health expenses >20% of income, and identified its predictors using multivariable logistic regression. All values were assessed as 2023 US$. Key cohorts were defined across strata with and without atherosclerotic cardiovascular disease (ASCVD) and/or heart failure (HF) before or up to 1 year after the T2D diagnosis.

**Results:** Overall, 106,881 patients with T2D followed for a median of 5.4 years (IQR: 3.1-7.5) had 2,258,376 healthcare visits, representing $3.56 billion in expected healthcare expenses. Annualized expected expenses among those without ASCVD/HF were $444 (147-4,471), compared with $2,930 (209-13,240) among those with ASCVD/HF. Across cohorts, 9-29% of patients with T2D had healthcare expenses above the threshold for financial hardship. Compared with White patients, Black and Hispanic patients were more likely, and Asians were less likely to have financial hardship (aOR: Black, 1.81 [1.73-1.91]; Hispanic, 1.39 [1.31-1.48]; Asian, 0.46 [0.37-0.57]).

**Conclusion:** A digital, individualized, expense-linked T2D registry showed that patients with (vs without) cardiovascular comorbidities had substantially higher medical expenses, with 1 in 5 facing financial hardship.

**GRAPHICAL ABSTRACT:** 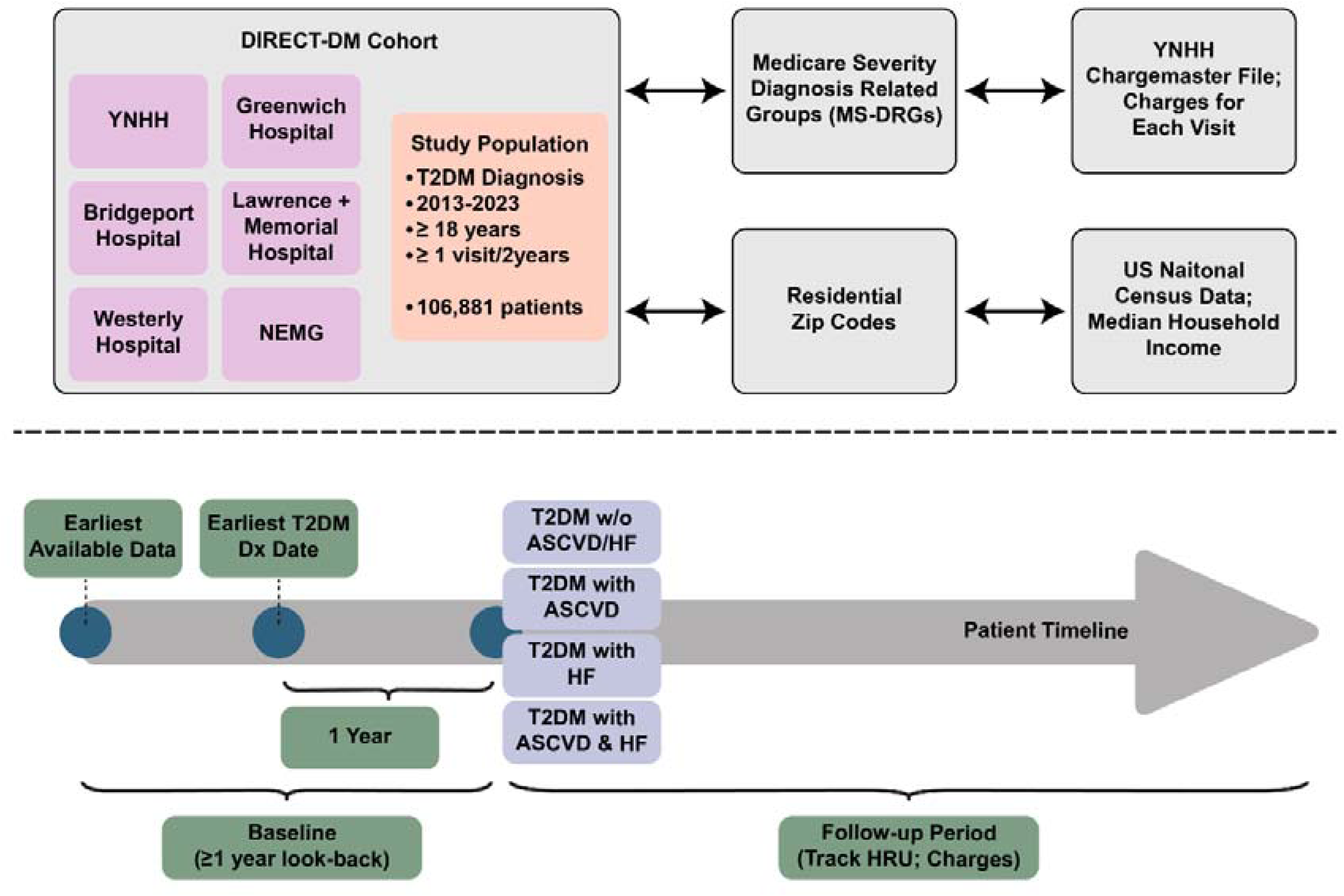

## INTRODUCTION

Despite the well-recognized financial burden that type 2 diabetes mellitus (T2D) imposes on patients, the assessment of real-world healthcare expenses remains fragmented and methodologically limited.^1,2^ Patients with T2D often have high healthcare resource utilization (HCRU), which can be exacerbated in the presence of cardiovascular comorbidities.^3^ Atherosclerotic cardiovascular disease (ASCVD) and heart failure (HF) are the most prevalent cardiovascular comorbidities in T2D and are associated with significantly high HCRU and medical expenses.^4,5^ These healthcare expenses can impose a substantial financial burden on patients, particularly for socioeconomically disadvantaged groups, who may be likely to experience financial hardship due to high out-of-pocket expenses.^6^ However, these approaches for evaluating healthcare expenses can be limited to self-selected cohorts or selected registries of patients with T2D.^7^

Traditional approaches for evaluating longitudinal medical expenses rely either on manual abstraction of data, which is resource-intensive and susceptible to selection bias, or on administrative claims-based analyses, which are often incomplete, outdated, and lack critical clinical granularity.^8,9^ These methodological limitations may lead to a skewed assessment of the healthcare utilization and financial challenges experienced by patients with T2D due to healthcare spending. However, recent government-mandated provisions on transparency in real-world hospital pricing under the 21^st^ Century Cures Act present an unprecedented opportunity to bridge these gaps by leveraging comprehensive electronic health record (EHR) data linked to real-world hospital charges.^10–12^

In this study, we conducted a longitudinal, system-wide assessment of HCRU and expenses in patients with T2D and cardiovascular comorbidities across a large and diverse US health system. Using retrospective EHR data integrated with real-world hospital charge information, this study provides a comprehensive, patient-centered evaluation of HCRU and the economic burden associated with T2D. This innovative methodology establishes a robust and scalable framework for evaluating real-world healthcare utilization and expenses.

## METHODS

The Yale Institutional Review Board approved the study protocol and waived the need for informed consent, as the study involves secondary analysis of pre-existing data.

### Data Source

We included data from the DIgital REgistry for CardiomeTabolic conditions – Diabetes Mellitus (DIRECT-DM) registry, a fully digital registry of patients with cardiometabolic conditions seeking care in the Yale New Haven Health System (YNHHS), with linkages to the US national census data and the Connecticut (CT) death index.^13^ The clinical data in the DIRECT-DM registry represent the EHRs from the YNHHS, a large, diverse, and integrated health system. The YNHHS includes (i) the Yale New Haven Hospital (YNHH), a 1,541-bed tertiary care center, and the primary teaching hospital for the Yale School of Medicine, in New Haven, (ii) the Bridgeport, Greenwich, Lawrence + Memorial, and Westerly Hospitals, representing 4 community hospitals, serving patients in Connecticut, New York, and Rhode Island, and (iii) the Northeast Medical Group (NEMG), an outpatient primary care network with over 130 practices across New England. All YNHHS sites use the Epic EHR system, and the clinical data in our study represent an extract from the Epic Clarity® database from January 1, 2013, to September 15, 2023.

### Cohort Creation

We identified all adult patients with a diagnosis of T2D across YNHHS using at least one International Classification of Diseases, Tenth Revision, Clinical Modification (ICD-10-CM) code for T2D (**Table S1**). Patients were categorized into four cohorts based on cardiovascular comorbidities (**Figure S1**). To define the study cohorts, we first determined the earliest recorded T2D diagnosis for each patient. We then assessed the presence of CVD by identifying relevant diagnoses within a one-year window before or after the T2D diagnosis. This approach ensured a minimum one-year look-back period to capture pre-existing CVD or exclusion criteria (**Table S2**) and a one-year follow-up period to confirm the presence of cardiovascular-related conditions. The study excluded patients with type 1 diabetes, gestational diabetes, or diabetes secondary to an underlying condition, a medication, or a chemical exposure, individuals with metastatic cancer, or organ transplants, those in hospice care, or diagnosed with HIV or end-stage renal disease. The index date, or cohort qualification date, was defined as the end of the one-year blanking period after the first T2D diagnosis. Based on the presence of the comorbid cardiovascular conditions before the index date, we categorized these patients into 4 cohorts: (i) those with T2D without ASCVD or HF, (ii) T2D and ASCVD alone, (iii) T2D with HF alone, or (iv) T2D with both ASCVD and HF.

### Healthcare Resource Utilization

To assess HCRU, we included only patients with one or more visits in YNHHS every 2 years until their death or end of follow-up. This strategy aimed to ensure an accurate assessment of HCRU by identifying a subset of patients who were receiving care continuously within the YNHHS system.

Across all 4 cohorts, these patients were followed longitudinally to evaluate patterns of healthcare utilization. HCRU was measured as the number of inpatient, outpatient, and emergency department (ED) visits. We calculated cumulative counts for patient visits across cohorts and annualized counts at the patient level.

We repeated these analyses for HCRU due to cardiovascular causes. For this, we defined the principal diagnosis codes for each visit, mapping the ICD-10 classification to the Clinical Classifications Software Refined (CCSR).^14^ Visits with CCSR codes starting with “CIR” were classified as those due to cardiovascular causes.

### Financial Burden of Care

Our approach for assessing financial burden of T2D care employed the Medicare severity diagnosis related groups (MS-DRGs) and linkages to the publicly available Chargemaster file.^15,16^ The MS-DRG classification system describes groupings of clinically similar conditions and procedures that are expected to require similar levels of hospital resources. Each hospital visit in YNHHS was classified into a specific MS-DRG which is used to assign a charge for the visit based on the identified diagnoses and procedures a patient underwent.^15,16^ We identified the primary diagnosis codes associated with encounters and included operating as well as non-operating room procedures. In case of multiple qualifying DRG codes, we selected the one with the higher weight, as is standard in reimbursement procedures.

Using the assigned DRG codes for each visit, we linked the encounter-level EHR data with the publicly available 2023 Yale chargemaster file. The 21st Century Cures Act made reporting of hospital-level service prices mandatory, dictating the availability of machine-readable chargemaster files.^17^ These files have information about the gross charges that are charged by the hospital without any discount, the discounted cash price applying to cash payers, and negotiated charges that are negotiated with Medicare or insurance providers. From the chargemaster file, we extracted Medicare and insurance-negotiated cash prices. Of note, we used the Medicare-negotiated cash prices for patients over 65 years of age at the time of visit, and the median insurance-negotiated cash prices for those under 65 years. All expenses were assessed as 2023 US dollars.

### Census Data Linkage and Financial Hardship

The YNHHS EHR included residential zip code information for all patients. Using these zip codes, we linked the EHR to the 2020 US national census data to estimate a patient’s household income, representing the median household income in their respective residential zip codes. We defined two categorical outcomes based on prior studies suggesting adverse financial outcomes if the healthcare expenses exceed certain thresholds of household income. These include (1) financial hardship, which is defined as annual healthcare expense greater than 20% of household income, and (2) catastrophic health expense, defined as an annual healthcare expense over 40% of household income.^6,18,19^ The latter is considered a financially ruinous expense and is used by the World Bank as a measure of extreme financial burden from healthcare expenses.^20,21^

### Statistical Analysis

Categorical variables are presented as frequency and percentage. Continuous variables are presented as mean and standard deviation or median and interquartile range (IQR), as appropriate. The comorbidity burden was reported using the Charlson Comorbidity Index, which ranges from 0 to 37, with higher scores indicating greater comorbidity burden. It includes the patient’s age and conditions such as myocardial infarction, HF, T2D, chronic pulmonary disease, and renal disease, each assigned a weighted score based on their severity and impact on mortality risk.^22,23^ These conditions were defined based on diagnosis codes before the index date (**Table S3**). Notably, among patients who died during the follow-up period, we also evaluated the HCRU and financial burden in the final year of life. Across cohorts, HCRU and financial burden were compared using analysis of variance (ANOVA) or Chi-square tests, as appropriate. All expenses were assessed as 2023 US dollars. Finally, we used multivariable logistic regression to identify factors associated with financial hardship. All statistical tests were 2-sided with a level of significance of 0.05. All analyses were performed using Python 3.9. Our study follows the Strengthening the Reporting of Observational Studies in Epidemiology (STROBE) reporting guidelines for observational cohort studies (**Table S4**).^24^

## RESULTS

### Study Population

We identified 223,932 patients in YNNHS with T2D from January 1, 2013, to September 15, 2023 (**Table S5**). We excluded patients who died (N = 8,574) or met any exclusion criteria on or before the cohort qualification date (N = 20,920), as well as those who did not seek regular care in YNHHS (N = 87,557). The final cohort had 106,881 patients with T2D, all of whom had at least one visit every two years from their cohort qualification date until either the cohort censor date (September 15, 2023) or their date of death. The median age of the study population was 62.8 (IQR, 52.4-72.6) years, with 59,460 (55.6%) women (**Table 1**). There were 64,650 (60.5%) who identified as non-Hispanic White, 19,326 (18.1%) as non-Hispanic Black, 2,398 (2.2%) as non-Hispanic Asian, and 16,825 (15.7%) as Hispanic, while 2,416 (2.3%) belonged to other race groups, and race/ethnicity data were missing for 1,266 (1.2%). A total of 15,220 (14.2%) of the patients were covered by commercial insurance providers, 47,961 (44.9%) by Medicare, 19,159 (17.9%) by Medicaid, 4,974 (4.7%) by managed care plans, and 19,567 (18.3%) were uninsured (**Table 1**).

**Table 1.** Study population characteristics.

| Characteristic of the Study Population | Patients with T2D | Patients with T2D without ASCVD or HF | Patients with T2D & ASCVD | Patients with T2D & HF | Patients with T2D & ASCVD & HF |
| --- | --- | --- | --- | --- | --- |
| **Number of patients** | 106,881 | 73,123 | 22,680 | 3,027 | 8,051 |
| **Demographics** |  |  |  |  |  |
| Age, Median (IQR) | 62.8 (52.4, 72.6) | 59.3 (49.2, 69.0) | 69.0 (60.2, 76.9) | 67.9 (57.3, 79.2) | 73.7 (64.0, 82.3) |
| Age Group, n (%) |  |  |  |  |  |
| 18-44 | 14,348 (13.4) | 13,087 (17.9) | 853 (3.8) | 229 (7.6) | 179 (2.2) |
| 45-64 | 44,986 (42.1) | 34,252 (46.8) | 7,663 (33.8) | 1,066 (35.2) | 2,005 (24.9) |
| ≥65 | 47,547 (44.5) | 25,784 (35.3) | 14,164 (62.5) | 1,732 (57.2) | 5,867 (72.9) |
| Sex |  |  |  |  |  |
| Female, n (%) | 59,460 (55.6) | 44,088 (60.3) | 10,100 (44.5) | 1,784 (58.9) | 3,488 (43.3) |
| Race, n (%) |  |  |  |  |  |
| White | 64,650 (60.5) | 41,142 (56.3) | 15,688 (69.2) | 1,979 (65.4) | 5,841 (72.5) |
| Black | 19,326 (18.1) | 14,285 (19.5) | 3,148 (13.9) | 615 (20.3) | 1,278 (15.9) |
| Hispanic | 16,825 (15.7) | 13,067 (17.9) | 2,743 (12.1) | 309 (10.2) | 706 (8.8) |
| Asian | 2,398 (2.2) | 1,903 (2.6) | 398 (1.8) | 31 (1.0) | 66 (0.8) |
| Other | 2,416 (2.3) | 1,783 (2.4) | 477 (2.1) | 51 (1.7) | 105 (1.3) |
| Missing | 1,266 (1.2) | 943 (1.3) | 226 (1.0) | 42 (1.4) | 55 (0.7) |
| **Health Insurance Plan Type, n (%)** |  |  |  |  |  |
| Commercial | 15,220 (14.2) | 12,589 (17.2) | 1,986 (8.8) | 225 (7.4) | 420 (5.2) |
| Medicare | 47,961 (44.9) | 28,041 (38.3) | 13,210 (58.2) | 1,652 (54.6) | 5,058 (62.8) |
| Medicaid | 19,159 (17.9) | 14,685 (20.1) | 3,016 (13.3) | 493 (16.3) | 965 (12.0) |
| Managed Care | 4,974 (4.7) | 4,134 (5.7) | 641 (2.8) | 74 (2.4) | 125 (1.6) |
| Uninsured | 19,567 (18.3) | 13,674 (18.7) | 3,827 (16.9) | 583 (19.3) | 1,483 (18.4) |
| **Clinical Comorbidities, n (%)** □ |  |  |  |  |  |
| CCI (Charlson Comorbidity Index) (Median, IQR) | 2 (1-3) | 1 (1-2) | 2 (2-4) | 3 (2-5) | 5 (3-6) |
| Myocardial Infarction | 4,714 (4.4) | 2 (0.0) | 2,532 (11.2) | 0 | 2,180 (27.1) |
| Congestive Heart Failure | 11,463 (10.7) | 117 (0.2) | 268 (1.2) | 3,027 (100.0) | 8,051 (100.0) |
| Peripheral Vascular Disease | 6,637 (6.2) | 117 (0.2) | 4,447 (19.6) | 18 (0.6) | 2,055 (25.5) |
| Cerebrovascular Accident or Transient Ischemic Attack | 9,621 (9.0) | 134 (0.2) | 7,052 (31.1) | 4 (0.1) | 2,431 (30.2) |
| Dementia | 2,292 (2.1) | 722 (1.0) | 787 (3.5) | 141 (4.7) | 642 (8.0) |
| Chronic Obstructive Pulmonary Disease | 21,456 (20.1) | 11,383 (15.6) | 5,080 (22.4) | 1,236 (40.8) | 3,757 (46.7) |
| Connective Tissue Disease | 4,063 (3.8) | 2,360 (3.2) | 1,029 (4.5) | 147 (4.9) | 527 (6.5) |
| Peptic Ulcer Disease | 1,951 (1.8) | 916 (1.3) | 556 (2.5) | 78 (2.6) | 401 (5.0) |
| Mild Liver Disease | 9,579 (9.0) | 6,358 (8.7) | 2,061 (9.1) | 297 (9.8) | 863 (10.7) |
| Uncomplicated Diabetes | 90,309 (84.5) | 64,678 (88.5) | 17,961 (79.2) | 2,310 (76.3) | 5,360 (66.6) |
| Hemiplegia | 1,353 (1.3) | 293 (0.4) | 712 (3.1) | 19 (0.6) | 329 (4.1) |
| Moderate to Severe Chronic Kidney Disease | 8,566 (8.0) | 3,082 (4.2) | 2,374 (10.5) | 586 (19.4) | 2,524 (31.4) |
| Diabetes with End-organ Damage | 1,6572 (15.5) | 8,445 (11.5) | 4,719 (20.8) | 717 (23.7) | 2,691 (33.4) |
| Localized Solid Tumor/Leukemia/Lymphoma | 10,411 (9.7) | 6,284 (8.6) | 2,675 (11.8) | 332 (11.0) | 1,120 (13.9) |
| Moderate to Severe Liver Disease | 851 (0.8) | 444 (0.6) | 190 (0.8) | 65 (2.1) | 152 (1.9) |
| Metastatic Solid Tumor | 0 | 0 | 0 | 0 | 0 |
| AIDS | 0 | 0 | 0 | 0 | 0 |

ASCVD, atherosclerotic cardiovascular disease; HF, heart failure; IQR, interquartile range; T2D, type-2 diabetes mellitus

There were 73,123 (68.4%) T2D patients without ASCVD or HF, 22,680 (21.2%) with ASCVD alone, 3,027 (2.8%) with HF alone and 8,051 (7.5%) with both ASCVD and HF (**Table 1**). Patients had a median CCI score of 2 (IQR, 1-3), ranging from a median of 1 (IQR, 1-2) in patients without ASCVD or HF to 5 (IQR, 3-6) in patients with both ASCVD and HF. A total of 9,587 (9.0%) patients died during the study period. Mortality rates varied across cohorts, ranging from 5.5% (4,020 individuals) in the cohort without ASCVD or HF to 25.8% (2,078 individuals) in those with T2D and both cardiovascular comorbidities (**Table S10**).

### Healthcare Utilization

Over a median follow-up of 5.4 (IQR, 3.1-7.5) years, the patients collectively experienced 147,919 inpatient, 250,151 ED, and 1,860,306 outpatient visits. During the study period, 43,387 (40.6%) patients had at least one inpatient admission, 55,662 (52.1%) visited the emergency department (ED), and 90,417 (84.6%) had at least one outpatient visit. Across the clinical cohorts, those with T2D without ASCVD or HF, the largest cohort, accounted for a majority of the visits, or 1,509,015 visits cumulatively. This was followed by patients with ASCVD alone, who had a total of 511,782 visits, while patients with HF alone had 69,829 visits and those with both ASCVD and HF had 167,750 visits (**Table S6**).

After accounting for differences in the number of patients and follow-up time, patients with T2D with both ASCVD and HF had the highest number of visits with a median 5.02 (IQR, 2.51-9.48) visits per patient per year, compared with 4.40 (IQR, 2.20-8.05) for those with HF, 3.74 (IQR, 1.92-6.80) for those with ASCVD, and 3.03 (IQR, 1.66-5.44) for patients without ASCVD or HF (**Table S6**). These results were consistent for inpatient, ED, and outpatient visits (**Table S6**), and similar patterns were also observed for visits due to cardiovascular causes (**Table S7**).

### Financial Burden of Care

Medical expenses represented a total of $3.6 billion across the study period. The cumulative healthcare expense was highest for the cohort of patients with T2D alone ($2.0 billion), which was driven by the size of this cohort (**Figure 1**). Patients with T2D and cardiovascular comorbidities had a total healthcare expense of over $1.6 billion, including $878.9 million for those with ASCVD alone, $201.4 million for those with HF alone, and $498.6 million for those with both ASCVD and HF (**Table 2**). Healthcare expenses for visits due to cardiovascular causes represented 16.3% ($323.9 million/$2.0 billion) of the total financial burden in those without ASCVD or HF, and 29.9% ($471.3 million/$1.6 billion) in those with ASCVD, HF, or both combined. The spending patterns across cohorts align with overall healthcare expenditures (**Table S8**).

**Figure 1.**
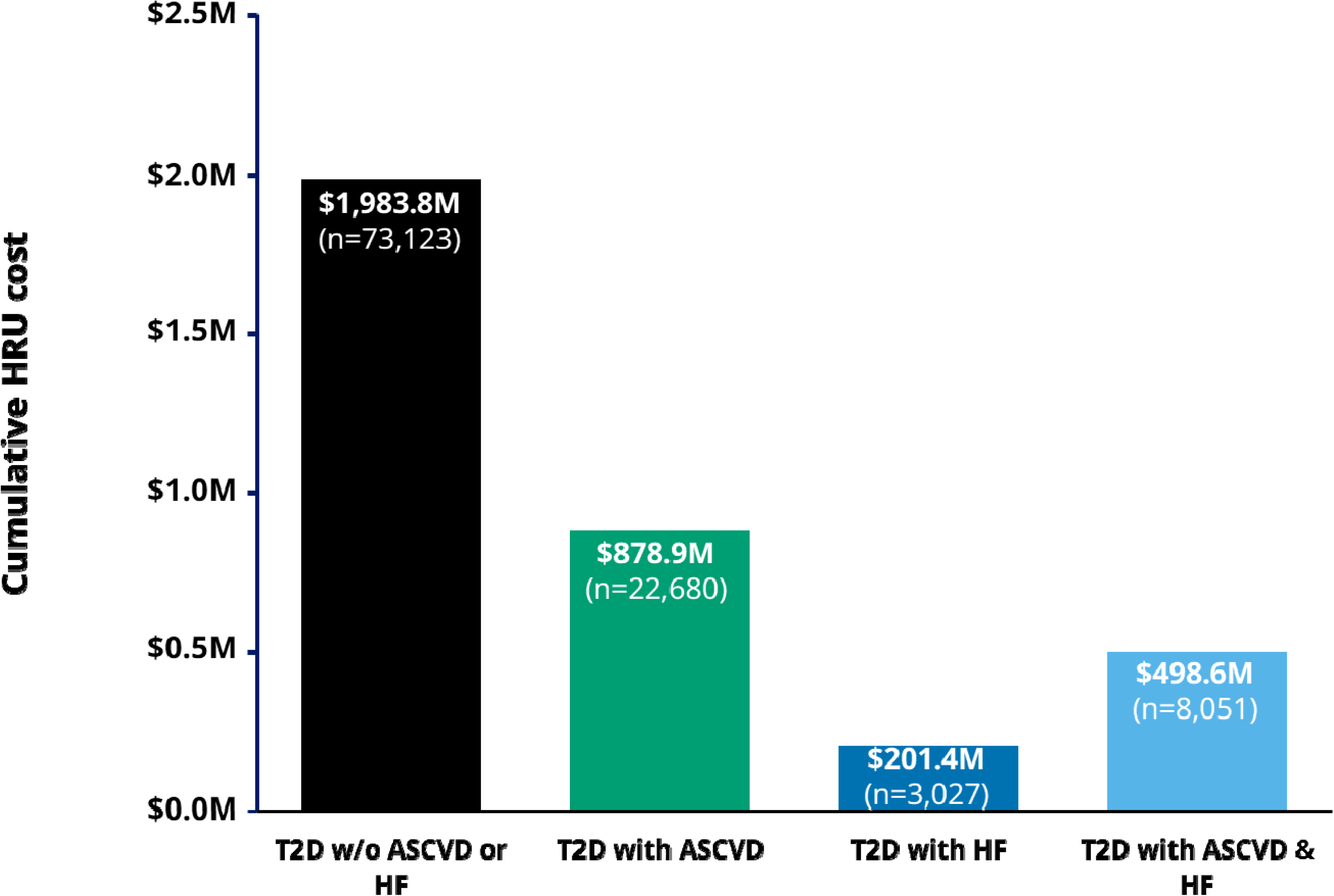
Cumulative overall healthcare expenses across study cohorts. Abbreviations: ASCVD, atherosclerotic cardiovascular disease; HF, heart failure; T2D, type-2 diabetes mellitus.

**Table 2.** Financial burden associated with healthcare resource utilization across study cohorts.

| Type of Visit | Metric | Overall Population<br>(N = 106,881) | T2D without<br>ASCVD or HF<br>(N = 73,123) | T2D with ASCVD &<br>HF<br>(N = 33,758) | T2D with ASCVD<br>(N = 22,680) | T2D with HF<br>(N = 3,027) | T2D with ASCVD<br>& HF<br>(N = 8,051) |
| --- | --- | --- | --- | --- | --- | --- | --- |
| All visits | Overall charges | \$3,562.7M | \$1,983.8M | \$1,578.9M | \$878.9M | \$201.4M | \$498.6M |
| | Total charges per patient,<br>Median (IQR) | \$1,792 (369-28,897) | \$1,476 (410-21,764) | \$7,531 (287-46,772) | \$3,248 (328-39,604) | \$13,714 (328-57,000) | \$16,046 (164-65,362) |
| | Annualized charges per<br>patient, Median (IQR) | \$653 (159-6,686) | \$444 (147-4,471) | \$2,930 (209-13,240) | \$1,783 (175-9,376) | \$4,791 (302-18,333) | \$8,336 (514-26,483) |
| Inpatient Visits | Overall charges | \$3,408.1M | \$1,871.6M | \$1,536.5M | \$849.5M | \$197.2M | \$489.9M |
| | Total charges per patient,<br>Median (IQR) | \$0 (0-27 178) | \$0 (0-20 202) | \$6,707 (0-45,050) | \$0 (0-38,089) | \$12,186 (0-55,384) | \$15,331 (0-64,143) |
| | Annualized charges per<br>patient, Median (IQR) | \$0 (0-6,336) | \$0 (0-4,110) | \$2,622 (0-12,878) | \$1,416 (0-9,062) | \$4,459 (0-17,849) | \$7,987 (0-26,084) |
| ED Visits | Overall charges | \$21.3M | \$15.9M | \$5.4M | \$3.7M | \$0.6M | \$1.2M |
| | Total charges per patient,<br>Median (IQR) | \$41 (0-164) | \$41 (0-224) | \$41 (0-123) | \$41 (0-123) | \$41 (0-153) | \$0 (0-123) |
| | Annualized charges per<br>patient, Median (IQR) | \$10 (0-42) | \$10 (0-44) | \$9 (0-37) | \$9 (0-35) | \$12 (0-44) | \$11 (0-44) |
| Outpatient Visits | Overall charges | \$133.3M | \$96.4M | \$36.9M | \$25.7M | \$3.7M | \$7.6M |
| | Total charges per patient,<br>Median (IQR) | \$656 (205-1,568) | \$697 (224-1,680) | \$492 (112-1,344) | \$560 (164-1,394) | \$492 (112-1,517) | \$328 (41-1,107) |
| | Annualized charges per<br>patient, Median (IQR) | \$170 (78-349) | \$174 (82-351) | \$160 (68-343) | \$158 (69-329) | \$174 (68-402) | \$163 (63-374) |

ASCVD, atherosclerotic cardiovascular disease; ED, emergency department; HF, heart failure; IQR, Interquartile Range; T2D, type-2 diabetes mellitus.

Overall, the median annualized expenses per patient were $653 (IQR, 159-6,686), with the highest annualized burden among patients with T2D with both ASCVD and HF ($8,336 [IQR, 514-26,483]) and lowest among those without cardiovascular comorbidities ($444 [IQR, 147-4,471]; **Table 2**). Of note, for patients with T2D without ASCVD or HF, the median charges from inpatient visits were $0 (IQR, 0-4,110), compared with $1,416 (IQR, 0-9,062) for those with ASCVD alone, $4,459 (IQR, 0-17,849) for those with HF alone, and $7,987 (IQR, 0-26,084) for those with both ASCVD and HF. For patients across all cohorts, the median annualized emergency and outpatient charges were below $50 and $200, respectively (**Table 2**).

### Financial Hardship and Catastrophic Healthcare Expenses

In the study population, 12,466 (11.7%) patients faced a financial obligation for healthcare greater than 20% of their annual household income. This was higher among those with a higher burden of CVD, with 8.6% of patients with T2D without ASCVD or HF at risk for financial hardship, compared with 14.0% with ASCVD, 23.6% with HF, and 28.7% with both ASCVD and HF (**Figure 3; Table S9**).

Furthermore, relative to White patients, Black and Hispanic patients were more likely, and Asians were less likely to have financial hardship (OR: Black, 1.81 [1.73-1.91]; Hispanic, 1.39 [1.31-1.48]; Asian, 0.46 [0.37-0.57]), after accounting for age, sex, and comorbidities. Comorbid HF at baseline was associated with a 2-fold higher odds of financial hardship (OR: HF, 2.20 [2.00-2.42]; ASCVD and HF, 1.98 [1.84-2.14]) compared with T2D alone (**Figure 2**).

**Figure 2.**
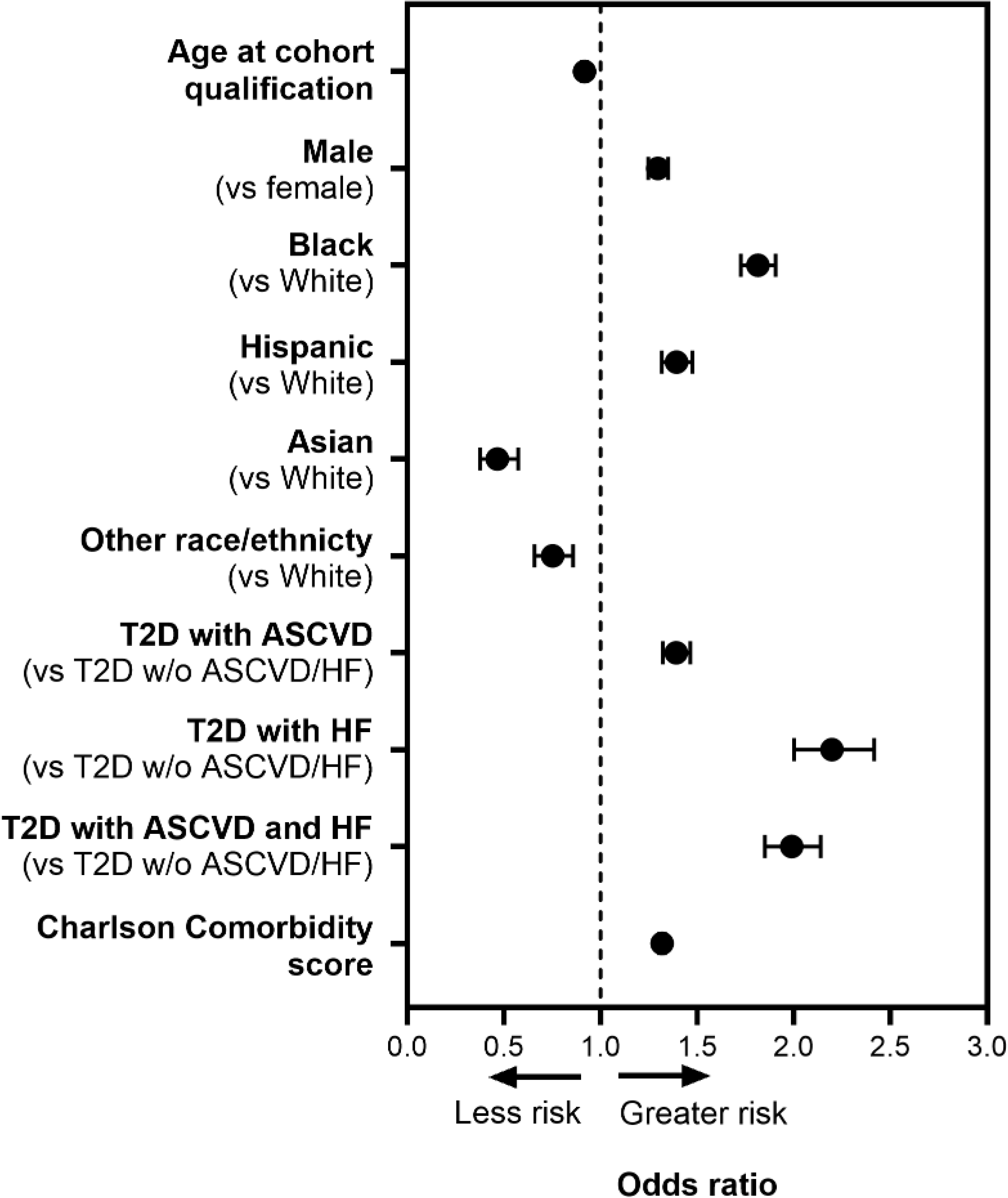
Factors associated with risk for financial hardship due to healthcare expenses. Abbreviations: ASCVD, atherosclerotic cardiovascular disease; HF, heart failure; T2D, type-2 diabetes mellitus.

Of note, 6,352 (5.9%) of patients had expenses that crossed the threshold for a catastrophic health expense, i.e. >40% of income (**Table S9**). The proportion was lowest among those with T2D alone (4.1%) and progressively higher for those with ASCVD (7.0%), HF (13.5%), and both ASCVD and HF (17.2%).

### Care in the Final Year of Life

Among 9,597 patients who died during the study follow-up, healthcare visits in the final year of life constituted 33.8% (90,702/268,183) of overall visits, including 50.2% (19,001/37,873) of all inpatient visits, 30.4% (7,212/23,691) of ED visits, and 31.2% (64,489/206,619) of outpatient visits (**Table S10**). Across cohorts, financial expenses in the final year of life constituted 52.5-55.3% of total healthcare expenses (**Table S10**). Further, across cohorts, a majority of patients were at a risk for financial hardship due to healthcare expenses in the final year of life (**Figure 3**).

**Figure 3.**
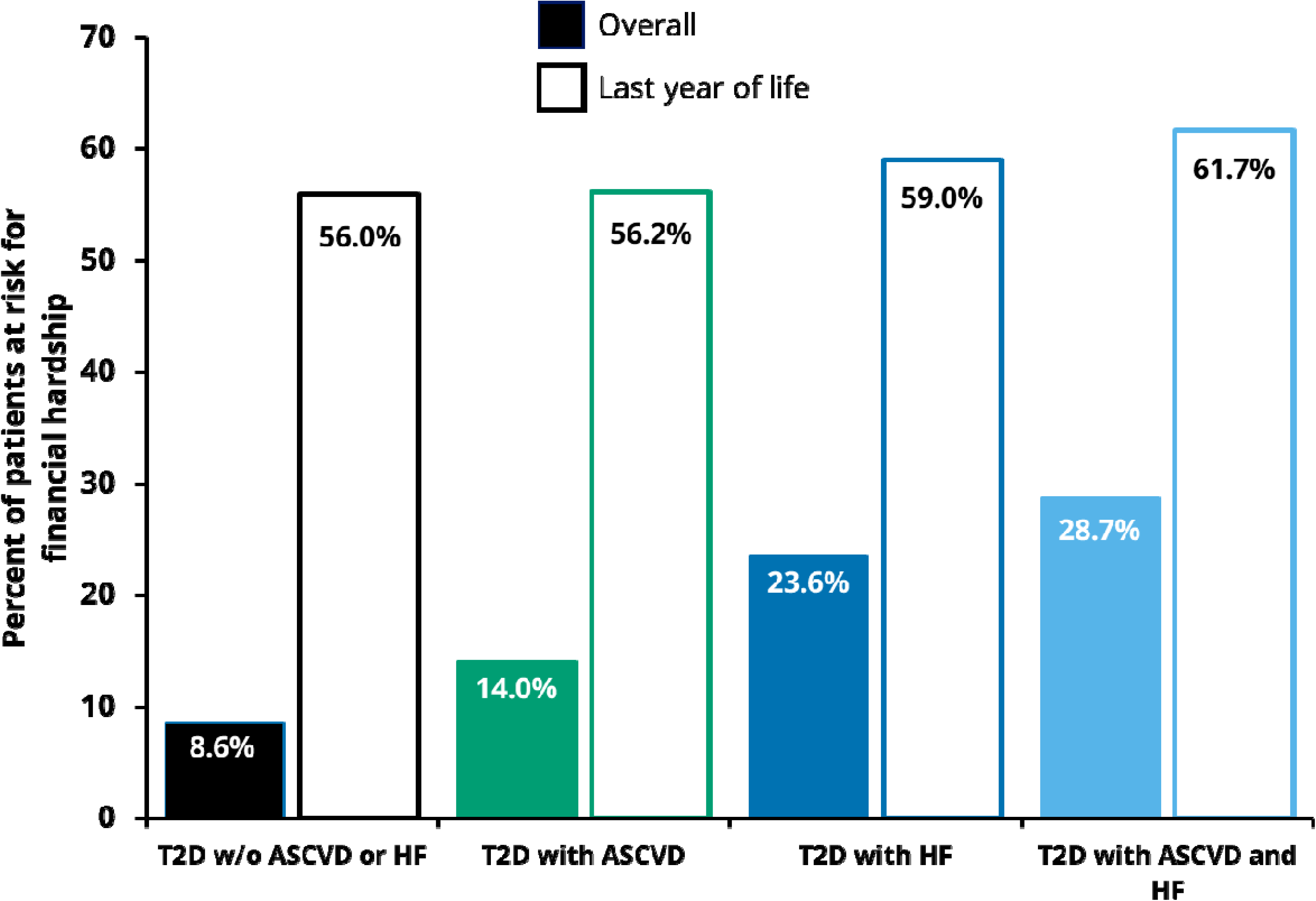
Proportion of people at risk for financial hardship due to healthcare expenses. Abbreviations: ASCVD, atherosclerotic cardiovascular disease; HF, heart failure; T2D, type-2 diabetes mellitus.

## DISCUSSION

In this study, we conducted a comprehensive, real-world assessment of HCRU and financial burden in patients with T2D without and with cardiovascular comorbidities across a large, integrated US health system. In this retrospective pragmatic cohort with a median follow-up of 5.4 years, and linked to hospital- and insurance-specific billing information, patients with T2D incurred more than $3.5 billion in healthcare expenses. Across cohorts, 9-29% of patients with T2D had healthcare expenses above the threshold for financial hardship. Further, 1 in 10 of those with T2DM and ASCVD and/or HF were at risk for a catastrophic healthcare expense, compared with 4% of those without cardiovascular comorbidities. There was a disproportionately higher burden of financially ruinous expenses among individuals from racial and ethnic minority groups. Of note, among those who died during follow-up, over half of the healthcare expenses were incurred in the final year of life. These findings highlight the high and inequitable financial hardship and toxicity associated with T2D, particularly in the presence of cardiovascular comorbidities.

Our results reinforce the disproportionate financial impact of CVD in patients with T2D.^1^ Healthcare utilization was highest in patients with both ASCVD and HF, who had the highest number of inpatient admissions as well as the highest annualized charges per patient. These patterns are consistent with prior research demonstrating that cardiovascular complications are among the primary drivers of morbidity, mortality, and cost in T2D.^1,25^ Importantly, our study extends these insights by quantifying the real-world financial burden of CVD within a diverse, multi-hospital health system, providing a more comprehensive view of expense distribution across different patient populations.

Beyond differences in utilization, we observed widespread financial hardship, particularly among those with cardiovascular comorbidities. Among patients with ASCVD and/or HF, nearly 1 in every 5, incurred expenses exceeding 20% of their household income. These findings build upon national surveys such as the National Health Interview Survey and the Medical Expenditure Panel Survey, which have documented high levels of financial distress among individuals with chronic disease.^26–28^ For example, nearly half of individuals with T2D and ASCVD reported financial difficulties, with nearly a quarter unable to pay their medical bills entirely.^27^ However, assessments relied primarily on interview-based surveys and population-level weighted estimates. In contrast, our analyses directly linked individual-level clinical and billing data, allowing for a precise and patient-centered assessment of real-world financial exposure. Additionally, our results highlight racial and ethnic disparities in financial burden, with Black and Hispanic patients being at significantly higher risk of financial hardship from health expenses, even after accounting for age, sex, and clinical comorbidities. These patterns underscore the need for equitable healthcare policies that address structural barriers to affordable, high-quality care.

Finally, a key contribution of our work is its demonstration of using real-world data and price transparency to assess health expenses at scale. The work leverages hospital chargemaster-derived expenses mandated by the 21^st^ Century Cures Act against clinical data from the EHR.^29^ The integrated expense-linked registry used in this study provides a scalable framework for real-world monitoring of healthcare expenses, allowing for tracking longitudinal trends, identifying high-risk populations, and implementing targeted interventions.^13^ Additionally, the ability to compare charges across hospitals using standardized pricing data unlocks opportunities for benchmarking and improving cost-efficiency within and across health systems.^30^

This study has limitations that must be considered when interpreting the results. First, this is a retrospective observational study, relying on EHR data, which may not be complete or may contain inaccuracies due to imperfect clinical and billing documentation. Our reliance on diagnosis codes to identify comorbidities and healthcare utilization, while standard in large-scale EHR studies, may be subject to misclassification.^31^ Nonetheless, we used validated coding systems and applied a one-year look-back and follow-up period to improve the accuracy of comorbidity identification. Additionally, our approach classified visits based on principal diagnosis codes mapped to the CCSR system, which has been widely used for standardizing diagnosis categories in research.^32^ Second, the study population may not represent the broader population with T2D and cardiovascular comorbidities, given the specific geographic and demographic characteristics of the YNHHS patient base. However, the YNHHS serves New Haven county, which is among the most representative of the US national population.^33,34^ Third, the charges associated with hospitalizations were determined based on the DRG codes assigned to each hospitalization and may not include all expenses incurred during the visit, such as charges for prescribed medications. However, DRG codes are used by the CMS to determine the charges of inpatient visits, and the differences from the actual expenses per visit are expected to be minor.^15^ Finally, although we used census-derived income estimates to assess financial hardship, household income at the ZIP-code level may not fully reflect individual-level financial circumstances. Among patients with comorbidities, financial hardship may be further underestimated, as household income could be impacted by reduced earning capacity due to illness-related work limitations. Future research should focus on advanced data science approaches, such as natural language processing, to extract additional information from unstructured EHR data, such as clinical notes and medical reports. These methods could improve the identification of comorbidities, healthcare encounters, and predictors of HCRU.

## CONCLUSION

In a digital, individualized, expense-linked registry of T2D patients enabled by recent US price transparency rules, we identified a substantial burden of healthcare expenses among patients with T2D, often exceeding their means. Specifically, one-fifth of patients with T2D and cardiovascular disease had expenses on healthcare exceeding 20% of their annual income, with those from racial/ethnic minority groups at highest risk for healthcare-related financial toxicity. Our approach can be leveraged for larger national assessments of the risk of financial harm from healthcare spending in T2D, to help identify those at risk of adverse financial outcomes as a result of their care needs.

## Data Availability

The data from the Yale New Haven Health System represent protected health information. To protect patient privacy, the Yale Institutional Review Board does not allow sharing of these data.

