## Supplementary material for "A Real-world Evaluation of Longitudinal Healthcare Expenses in a Health System Registry of Type-2 Diabetes Mellitus and Cardiovascular Disease Enabled by the 21st Century Cures Act": Online Supplement

### SUPPLEMENTAL MATERIALS

**Figure S1. Cohort qualification methodology.** Abbreviations: ASCVD, atherosclerotic cardiovascular disease; HF, heart failure; T2DM, type-2 diabetes mellitus**.**

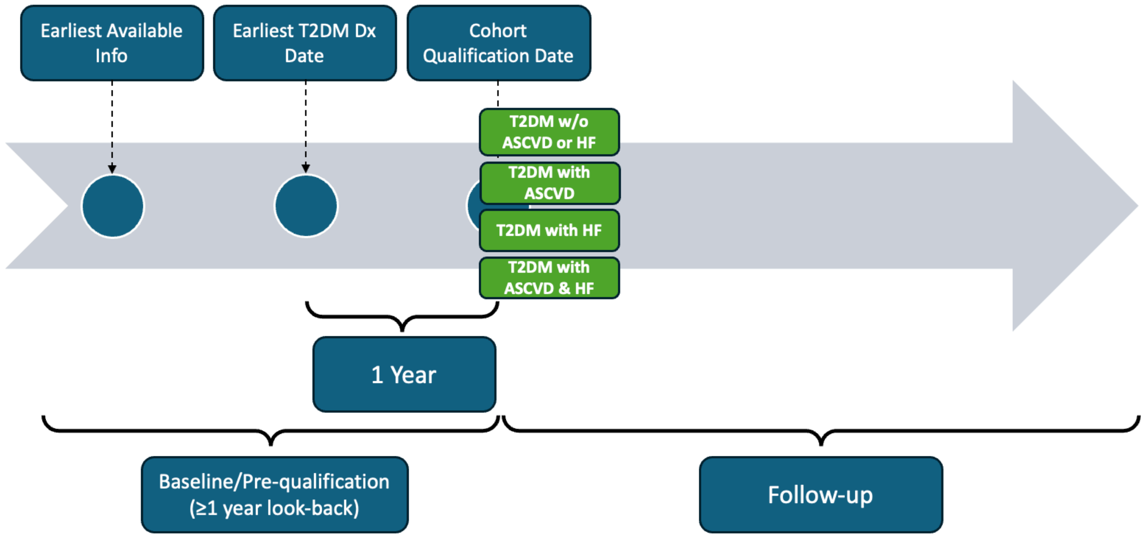

**Table S1. International Classification of Disease Tenth Revision codes for key conditions.** Abbreviations: ASCVD, atherosclerotic cardiovascular disease; HF, heart failure; T2DM, type-2 diabetes mellitus.

| **Condition** | | | **International Classification of Disease tenth revision (ICD-10) Codes** |
|---|---|---|---|
| **Heart Failure (HF)** | **Heart failure with preserved ejection fraction (HFpEF)** | | 'I5030', 'I5031', 'I5032', 'I5033' |
| | **Heart failure with reduced ejection fraction (HfrEF)** | | 'I5020', 'I5021', 'I5022' |
| | **Combined HfpEF and HfrEF** | | 'I5040', 'I5041', 'I5042', 'I5043' |
| | **Unspecified** | | 'I0981', 'I110', 'I130', 'I501', 'I5023', 'I50810', 'I50811', 'I50812', 'I50813', 'I50814', 'I5082', 'I5083', 'I5084', 'I5089', 'I509' |
| **Atherosclerotic Cardiovascular Disease (ASCVD)** | **Coronary heart disease (CHD)** | **Angina** | 'I200', 'I201', 'I202', 'I208', 'I209', 'I237', 'I25110', 'I25111', 'I25112', 'I25118', 'I25119', 'I25700', 'I25701', 'I25702', 'I25708', 'I25709', 'I25710', 'I25711', 'I25712', 'I25718', 'I25719', 'I25720', 'I25721', 'I25722', 'I25728', 'I25729', 'I25730', 'I25731', 'I25732', 'I25738', 'I25739', 'I25750', 'I25751', 'I25752', 'I25758', 'I25759', 'I25760', 'I25761', 'I25762', 'I25768', 'I25769', 'I25790', 'I25791', 'I25792', 'I25798', 'I25799', 'I25810' |
| | | **Myocardial infarction (MI)** | 'I2101', 'I2102', 'I2109', 'I2111', 'I2119', 'I2121', 'I2129', 'I213', 'I214', 'I219', 'I21A1', 'I21A9', 'I220', 'I221', 'I222', 'I228', 'I229', 'I230', 'I231', 'I232', 'I233', 'I234', 'I235', 'I236', 'I237', 'I238' |
| | | **Other acute and chronic ischemic heart disease** | 'I240', 'I241', 'I248', 'I249', 'I2510','I25110', 'I25111', 'I25112', 'I25118', 'I25119', 'I252', 'I253', 'I2541', 'I2542', 'I255', 'I256', 'I25700', 'I25701', 'I25702', 'I25708', 'I25709', 'I25710', 'I25711', 'I25712', 'I25718', 'I25719', 'I25720', 'I25721', 'I25722', 'I25728', 'I25729', 'I25730', 'I25731', 'I25732', 'I25738', 'I25739', 'I25750', 'I25751', 'I25752', 'I25758', 'I25759', 'I25760', 'I25761', 'I25762', 'I25768', 'I25769', 'I25790', 'I25791', 'I25792', 'I25798', 'I25799', 'I25810', 'I25811', 'I25812', 'I2582', 'I2583', 'I2584', 'I2589', 'I259' |
| | **Cerebrovascular disease** | **Transient ischemic attack (TIA)** | 'G450', 'G451', 'G452', 'G453', 'G454', 'G458', 'G459' |
| | | **Ischemic stroke** | 'G43601', 'G43609', 'G43611', 'G43619', 'I6300', 'I63011', 'I63012', 'I63013', 'I63019', 'I6302', 'I63031', 'I63032', 'I63033', 'I63039', 'I6309', 'I6310', 'I63111', 'I63112', 'I63113', 'I63119', 'I6312', 'I63131', 'I63132', 'I63133', 'I63139', 'I6319', 'I6320', 'I63211', 'I63212', 'I63213', 'I63219', 'I6322', 'I63231', 'I63232', 'I63233', 'I63239', 'I6329', 'I6330', 'I63311', 'I63312', 'I63313', 'I63319', 'I63321', 'I63322', 'I63323', 'I63329', 'I63331', 'I63332', 'I63333', 'I63339', 'I63341', 'I63342', 'I63343', 'I63349', 'I6339', 'I6340', 'I63411', 'I63412', 'I63413', 'I63419', 'I63421', 'I63422', 'I63423', 'I63429', 'I63431', 'I63432', 'I63433', 'I63439', 'I63441', 'I63442', 'I63443', 'I63449', |

| | | | |
|---|---|---|---|
| | | | 'I6349', 'I6350', 'I63511', 'I63512', 'I63513', 'I63519', 'I63521', 'I63522', 'I63523', 'I63529', 'I63531', 'I63532', 'I63533', 'I63539', 'I63541', 'I63542' 'I63543', 'I63549', 'I6359', 'I636', 'I638', 'I6381', 'I6389', 'I639', 'I6501', 'I6502', 'I6503', 'I6509', 'I651', 'I6521', 'I6522', 'I6523', 'I6529', 'I658', 'I659', 'I6601', 'I6602', 'I6603', 'I6609', 'I6611', 'I6612', 'I6613', 'I6619', 'I6621', 'I6622', 'I6623', 'I6629', 'I663', 'I668', 'I669', 'I672', 'I6781', 'I6782', 'I6930' 'I6931', 'I69310', 'I69311', 'I69312', 'I69313', 'I69314', 'I69315', 'I69318', 'I69319', 'I69320', 'I69321' 'I69322', 'I69323', 'I69328', 'I69331', 'I69332', 'I69333', 'I69334', 'I69339', 'I69341', 'I69342', 'I69343', 'I69344', 'I69349', 'I69351', 'I69352', 'I69353', 'I69354', 'I69359', 'I69361', 'I69362', 'I69363', 'I69364', 'I69365', 'I69369', 'I69390', 'I69391', 'I69392', 'I69393', 'I69398', 'I6980', 'I6981', 'I69810', 'I69811', 'I69812', 'I69813', 'I69814', 'I69815', 'I69818', 'I69819', 'I69820', 'I69821', 'I69822', 'I69823', 'I69828', 'I69831', 'I69832', 'I69833', 'I69834', 'I69839', 'I69841', 'I69842', 'I69843', 'I69844', 'I69849', 'I69851', 'I69852', 'I69853', 'I69854', 'I69859', 'I69861', 'I69862', 'I69863', 'I69864', 'I69865', 'I69869', 'I69890', 'I69891', 'I69892', 'I69893', 'I69898', 'I6990', 'I6991', 'I69910', 'I69911', 'I69912', 'I69913', 'I69914', 'I69915', 'I69918', 'I69919', 'I69920', 'I69921', 'I69922', 'I69923', 'I69928', 'I69931', 'I69932', 'I69933', 'I69934', 'I69939', 'I69941', 'I69942', 'I69943', 'I69944', 'I69949', 'I69951', 'I69952', 'I69953', 'I69954', 'I69959', 'I69961', 'I69962', 'I69963', 'I69964', 'I69965', 'I69969', 'I69990', 'I69991', 'I69992', 'I69993', 'I69998' |
| | **Peripheral artery disease (PAD)** | | 'I70201', 'I70202', 'I70203', 'I70208', 'I70209', 'I70211', 'I70212', 'I70213', 'I70218', 'I70219', 'I70221', 'I70222', 'I70223', 'I70228', 'I70229', 'I70231', 'I70232', 'I70233', 'I70234', 'I70235', 'I70238', 'I70239', 'I70241', 'I70242', 'I70243', 'I70244', 'I70245', 'I70248', 'I70249', 'I7025', 'I70261', 'I70262', 'I70263', 'I70268', 'I70269', 'I70291', 'I70292', 'I70293', 'I70298' ,'I70299', 'I70301', 'I70302', 'I70303', 'I70308', 'I70309', 'I70311', 'I70312', 'I70313', 'I70318', 'I70319', 'I70321', 'I70322', 'I70323', 'I70328', 'I70329' ,'I70331', 'I70332', 'I70333', 'I70334', 'I70335', 'I70338', 'I70339', 'I70341', 'I70342', 'I70343', 'I70344', 'I70345', 'I70348', 'I70349', 'I7035' 'I70361', 'I70362', 'I70363', 'I70368', 'I70369', 'I70391', 'I70392', 'I70393', 'I70398', 'I70399', 'I70401', 'I70402', 'I70403', 'I70408', 'I70409', 'I70411', 'I70412', 'I70413', 'I70418', 'I70419', 'I70421', 'I70422', 'I70423', 'I70428', 'I70429', 'I70431', 'I70432', 'I70433', 'I70434', 'I70435', 'I70438', 'I70439', 'I70441', 'I70442', 'I70443', 'I70444', 'I70445', 'I70448', 'I70449', 'I7045', 'I70461', 'I70462', 'I70463', 'I70468', 'I70469', 'I70491', 'I70492', 'I70493', 'I70498', 'I70499', 'I70501', 'I70502', 'I70503', 'I70508', 'I70509', 'I70511', 'I70512', 'I70513', 'I70518', 'I70519', 'I70521', 'I70522', 'I70523', 'I70528', 'I70529', 'I70531', 'I70532', 'I70533', 'I70534', 'I70535', 'I70538', 'I70539', 'I70541', 'I70542', |

| | | |
|---|---|---|
| | | 'I70543', 'I70544', 'I70545', 'I70548', 'I70549', 'I7055', 'I70561', 'I70562', 'I70563', 'I70568', 'I70569', 'I70591', 'I70592', 'I70593', 'I70598', 'I70599', 'I70601', 'I70602', 'I70603', 'I70608', 'I70609', 'I70611', 'I70612', 'I70613', 'I70618', 'I70619', 'I70621', 'I70622', 'I70623', 'I70628', 'I70629', 'I70631', 'I70632', 'I70633', 'I70634', 'I70635', 'I70638', 'I70639', 'I70641', 'I70642', 'I70643', 'I70644', 'I70645', 'I70648', 'I70649', 'I7065', 'I70661', 'I70662', 'I70663', 'I70668', 'I70669', 'I70691', 'I70692', 'I70693', 'I70698', 'I70699', 'I70701', 'I70702', 'I70703', 'I70708', 'I70709', 'I70711', 'I70712', 'I70713', 'I70718', 'I70719', 'I70721', 'I70722', 'I70723', 'I70728', 'I70729', 'I70731', 'I70732', 'I70733', 'I70734', 'I70735', 'I70738', 'I70739', 'I70741', 'I70742', 'I70743', 'I70744', 'I70745', 'I70748', 'I70749', 'I7075', 'I70761', 'I70762', 'I70763', 'I70768', 'I70769', 'I70791', 'I70792', 'I70793', 'I70798', 'I70799', 'I7092', 'I739', 'I742', 'I743', 'I744', 'I745', 'I748', 'I749', 'I75011', 'I75012', 'I75013', 'I75019', 'I75021', 'I75022', 'I75023', 'I75029', 'I7581' 'I7589' |
| **Type 2 Diabetes Mellitus (T2D)** | | 'E1100', 'E1101', 'E1110', 'E1111', 'E1121', 'E1122', 'E1129', 'E11311', 'E11319', 'E11321', 'E113211', 'E113212', 'E113213', 'E113219', 'E11329', 'E113291', 'E113292', 'E113293', 'E113299', 'E11331', 'E113311', 'E113312', 'E113313', 'E113319', 'E11339', 'E113391', 'E113392', 'E113393', 'E113399', 'E11341', 'E113411', 'E113412', 'E113413', 'E113419', 'E11349', 'E113491', 'E113492', 'E113493', 'E113499', 'E11351', 'E113511', 'E113512', 'E113513', 'E113519', 'E113521', 'E113522', 'E113523', 'E113529', 'E113531', 'E113532', 'E113533', 'E113539', 'E113541', 'E113542', 'E113543', 'E113549', 'E113551', 'E113552', 'E113553', 'E113559', 'E11359', 'E113591', 'E113592' 'E113593', 'E113599', 'E1136', 'E1137X1', 'E1137X2', 'E1137X3', 'E1137X9', 'E1139', 'E1140', 'E1141', 'E1142', 'E1143', 'E1144', 'E1149', 'E1151', 'E1152', 'E1159', 'E11610', 'E11618', 'E11620', 'E11621', 'E11622', 'E11628', 'E11630', 'E11638', 'E11641', 'E11649', 'E1165', 'E1169', 'E118', 'E119' |

**Table S2. International Classification of Disease Tenth Revision Codes for the Identification of Exclusion Criteria Diagnoses.** Abbreviations: ASCVD, atherosclerotic cardiovascular disease; HF, heart failure; T2DM, type-2 diabetes mellitus.

| **Condition** | **International Classification of Disease Tenth Revision (ICD-10) Codes** |
|---|---|
| **Metastatic Cancer** | 'C77.x', 'C78.x', 'C79.x', 'C80.x' |
| **Gestational Diabetes** | 'O24.4', 'O24.41', 'O24.410', 'O24.414', 'O24.415', 'O24.419', 'O24.42', 'O24.420', 'O24.424', 'O24.425', 'O24.429', 'O24.43', 'O24.430', 'O24.434', 'O24.435', 'O24.439', 'O24.8', 'O24.81', 'O24.811', 'O24.812', 'O24.813', 'O24.819', 'O24.82', 'O24.83', 'O24.9', 'O24.91', 'O24.911', 'O24.912', 'O24.913', 'O24.919', 'O24.92', 'O24.93' |
| **Diabetes Mellitus due to Underlying Condition** | 'E08.x' |
| **Drug or Chemical Induced Diabetes Mellitus** | 'E09.x' |
| **Type 1 Diabetes Mellitus** | 'E10.x' |
| **Organ Transplant** | 'Z94.0', 'Z94.1', 'Z94.2', 'Z94.3', 'Z94.4', 'Z94.5', 'Z94.6', 'Z94.7', 'Z94.8', 'Z94.9' |
| **Hospice Care** | 'Z51.5' |
| **Human Immunodeficiency Virus** | 'B20.x','B21.x', 'B22.x', 'B24.x' |
| **End Stage Renal Disease** | 'N18.6', 'Z99.2' |

**Table S3. International Classification of Disease Tenth Revision Codes for the Identification of baseline comorbidities.**

| Condition | International Classification of Disease Tenth Revision (ICD-10) Codes | Weight in Charlson Comorbidity Index (CCI) |
|---|---|---|
| **Myocardial Infarction** | (See Table S1) | 1 |
| **Heart Failure** | (See Table S1) | 1 |
| **Peripheral Vascular Disease** | (See Table S1) | 1 |
| **Cerebrovascular Disease** | (See Table S1) | 1 |
| **Dementia** | 'F00.x', 'F01.x', 'F02.x', 'F03.x', 'F05.1x', 'G30.x', 'G31.1x' | 1 |
| **Chronic Pulmonary Disease** | 'I27.x', 'J40.x', 'J41.x', 'J42.x', 'J43.x', 'J44.x', 'J45.x', 'J46.x', 'J47.x', 'J60.x', 'J61.x', 'J62.x', 'J63.x', 'J64.x', 'J65.x', 'J66.x', 'J67.x', 'J68.x', 'J70.x' | 1 |
| **Connective Tissue Disease/Rheumatic Disease** | 'M05.x', 'M06.x', 'M31.x', 'M32.x', 'M33.x', 'M34.x', 'M35.x', 'M36.x' | 1 |
| **Peptic Ulcer Disease** | 'K25.x', 'K26.x', 'K27.x', 'K28.x' | 1 |
| **Mild Liver Disease** | 'B18.x', 'K70.0x', 'K70.1x', 'K70.2x', 'K70.3x', 'K70.9x', 'K71.3x', 'K71.4x', 'K71.5x', 'K71.7x', 'K73.x', 'K74.x', 'K76.0x', 'K76.2x', 'K76.3x', 'K76.4x', 'K76.8x', 'K76.9x', 'Z94.4x' | 1 |
| **Paraplegia/Hemiplegia** | 'G80.x', 'G81.x', 'G82.x', 'G83.x', 'G84.x', 'G04.1x', 'G11.4x' | 2 |
| **Renal Disease** | 'N03.x', 'N05.x', 'N18.x', 'N19.x', 'Z49.x', 'I12.0x', 'I13.11x', 'I13.2x', 'N25.0x', 'Z94.0x', 'Z99.2x' | 2 |
| **Cancer** | 'C00.x', 'C01.x', 'C02.x', 'C03.x', 'C04.x', 'C05.x', 'C06.x', 'C07.x', 'C08.x', 'C09.x', 'C10.x', 'C11.x', 'C12.x', 'C13.x', 'C14.x', 'C15.x', 'C16.x', 'C17.x', 'C18.x', 'C19.x', 'C20.x', 'C21.x', 'C22.x', 'C23.x', 'C24.x', 'C25.x', 'C26.x', 'C30.x', 'C31.x', 'C32.x', 'C33.x', 'C34.x', 'C37.x', 'C38.x', 'C39.x', 'C40.x', 'C41.x', 'C43.x', 'C45.x', 'C46.x', 'C47.x', 'C48.x', 'C49.x', 'C50.x', 'C51.x', 'C52.x', 'C53.x', 'C54.x', 'C55.x', 'C56.x', 'C57.x', 'C58.x', 'C60.x', 'C61.x', 'C62.x', 'C63.x', 'C64.x', 'C65.x', 'C66.x', 'C67.x', 'C68.x', 'C69.x', 'C70.x', 'C71.x', 'C72.x', 'C73.x', 'C74.x', 'C75.x', 'C76.x', 'C81.x', 'C82.x', 'C83.x', 'C84.x', 'C85.x', 'C88.x', 'C90.x', 'C91.x', 'C92.x', 'C93.x', 'C94.x', 'C95.x', 'C96.x', 'C97.x' | 2 |
| **Moderate or Severe Liver Disease** | 'I85.0x', 'I85.9x', 'I86.4x', 'I98.2x', 'K70.4x', 'K71.1x', 'K72.1x', 'K72.9x', 'K76.5x', 'K76.6x', 'K76.7x' | 3 |
| **Metastatic Carcinoma** | (See Table S2) | 6 |
| **HIV/AIDS** | (See Table S2) | 6 |

**Table S4. Checklist for the Strengthening the Reporting of Observational Studies in Epidemiology (STROBE) reporting guidelines.** Abbreviations: ASCVD, atherosclerotic cardiovascular disease; HF, heart failure; T2DM, type-2 diabetes mellitus.

| | Item No | Recommendation | Page No |
|---|---|---|---|
| **Title and abstract** | 1 | (*a*) Indicate the study's design with a commonly used term in the title or the abstract | 2 |
| | | (*b*) Provide in the abstract an informative and balanced summary of what was done and what was found | 2 |
| **Introduction** | | | |
| Background/rationale | 2 | Explain the scientific background and rationale for the investigation being reported | 3 |
| Objectives | 3 | State specific objectives, including any prespecified hypotheses | 3,4 |
| **Methods** | | | |
| Study design | 4 | Present key elements of study design early in the paper | 4,5 |
| Setting | 5 | Describe the setting, locations, and relevant dates, including periods of recruitment, exposure, follow-up, and data collection | 4,5 |
| Participants | 6 | (*a*) Give the eligibility criteria, and the sources and methods of selection of participants. Describe methods of follow-up | 4,5,Supplement |
| | | (*b*) For matched studies, give matching criteria and number of exposed and unexposed | NA |
| Variables | 7 | Clearly define all outcomes, exposures, predictors, potential confounders, and effect modifiers. Give diagnostic criteria, if applicable | 5-7 |
| Data sources/ measurement | 8* | For each variable of interest, give sources of data and details of methods of assessment (measurement). Describe comparability of assessment methods if there is more than one group | 5-7 |
| Bias | 9 | Describe any efforts to address potential sources of bias | NA |
| Study size | 10 | Explain how the study size was arrived at | 4,5 |
| Quantitative variables | 11 | Explain how quantitative variables were handled in the analyses. If applicable, describe which groupings were chosen and why | 5-7 |
| Statistical methods | 12 | (*a*) Describe all statistical methods, including those used to control for confounding | 6-8 |
| | | (*b*) Describe any methods used to examine subgroups and interactions | 7 |
| | | (*c*) Explain how missing data were addressed | 5-7 |
| | | (*d*) If applicable, explain how loss to follow-up was addressed | 5-7 |
| | | (*e*) Describe any sensitivity analyses | 7 |
| **Results** | | | |
| Participants | 13* | (a) Report numbers of individuals at each stage of study—eg numbers potentially eligible, examined for eligibility, confirmed eligible, included in the study, completing follow-up, and analysed | 8 |
| | | (b) Give reasons for non-participation at each stage | 8 |
| | | (c) Consider use of a flow diagram | Supplement |
| Descriptive data | 14* | (a) Give characteristics of study participants (eg demographic, clinical, social) and information on exposures and potential confounders | 8 |

| | | | |
|---|---|---|---|
| | | (b) Indicate number of participants with missing data for each variable of interest | NA |
| | | (c) Summarise follow-up time (eg, average and total amount) | 8 |
| Outcome data | 15* | Report numbers of outcome events or summary measures over time | 8-10 |

**Table S5. Study flow diagram.**

| Step | Criteria | N |
|---|---|---|
| 1 | Patients who were not research opt-out and are with T2D at the Yale New Haven Healthcare System (YNNHS) from January 1, 2013 to September 15, 2023 | 223,932 |
| 2 | From Step 1, excluded patient who died before cohort qualification date (within 1 year of earliest T2DM diagnosis) | 215,358 |
| 3 | From Step 2, excluded patients with diagnoses of cancer, end stage renal disease, HIV, organ transplant, T1DM, Diabetes Mellitus due to Underlying Condition, Drug or Chemical Induced Diabetes Mellitus, and those receiving hospice care on or before cohort qualification date | 194,438 |
| 4 | From step 3, included patients with at least 1 visit after cohort qualification date | 141,588 |
| 5 | From Step 4, included patients with at least 1 visit every 2 years from cohort qualification date to death or September 15, 2023 | 106,881 |

**Table S6. Healthcare resource utilization across study cohorts.** Abbreviations: ASCVD, atherosclerotic cardiovascular disease; HF, heart failure; T2DM, type-2 diabetes mellitus.

| Type of Visit | Metric | Overall Population (N = 106,881) | T2DM w/o ASCVD & HF (N = 73,123) | T2DM with ASCVD or HF (N = 33,758) | T2DM with ASCVD (N = 22,680) | T2DM with HF (N = 3,027) | T2DM with ASCVD & HF (N = 8,051) |
|---|---|---|---|---|---|---|---|
| **All visits** | **Total visits** | 2258376 | 1509015 | 749361 | 511782 | 69829 | 167750 |
| | **Total visits per patient (Median [IQR])** | 13 (4-27) | 13 (5-27) | 13 (4-29) | 13 (4-30) | 13 (3-30) | 11 (2-27) |
| | **Annualized visits per patient (Median [IQR])** | 3.3 (1.75-6.03) | 3.03 (1.66-5.44) | 4.05 (2.05-7.48) | 3.74 (1.92-6.8) | 4.4 (2.2-8.05) | 5.02 (2.51-9.48) |
| **Inpatient Visits** | **Total visits** | 147919 | 78286 | 69633 | 39333 | 7826 | 22474 |
| | **Total visits per patient (Median [IQR])** | 0 (0-1) | 0 (0-1) | 1 (0-3) | 0 (0-2) | 1 (0-3) | 1 (0-4) |
| | **Annualized visits per patient (Median [IQR])** | 0.0 (0.0-0.34) | 0.0 (0.0-0.23) | 0.19 (0.0-0.72) | 0.13 (0.0-0.51) | 0.28 (0.0-0.93) | 0.5 (0.0-1.49) |
| **ED Visits** | **Total visits** | 250151 | 174247 | 75904 | 51511 | 7356 | 17037 |
| | **Total visits per patient (Median [IQR])** | 1 (0-3) | 1 (0-3) | 1 (0-2) | 1 (0-2) | 1 (0-3) | 0 (0-2) |
| | **Annualized visits per patient (Median [IQR])** | 0.16 (0.0-0.6) | 0.15 (0.0-0.56) | 0.19 (0.0-0.67) | 0.17 (0.0-0.61) | 0.22 (0.0-0.75) | 0.24 (0.0-0.83) |
| **Outpatient Visits** | **Total visits** | 1860306 | 1256482 | 603824 | 420938 | 54647 | 128239 |
| | **Total visits per patient (Median [IQR])** | 10 (3-23) | 10 (3-22) | 9 (2-23) | 10 (3-24) | 9 (2-24) | 7 (1-20) |
| | **Annualized visits per patient (Median [IQR])** | 2.58 (1.26-4.91) | 2.45 (1.23-4.53) | 2.95 (1.33-5.84) | 2.89 (1.35-5.55) | 2.96 (1.27-6.07) | 3.22 (1.33-6.82) |

**Table S7. Healthcare resource utilization due to cardiovascular causes across study cohorts.** Abbreviations: ASCVD, atherosclerotic cardiovascular disease; HF, heart failure; T2DM, type-2 diabetes mellitus.

| Type of Visit | Metric | Overall Population (N = 106,881) | T2DM w/o ASCVD & HF (N = 73,123) | T2DM with ASCVD or HF (N = 33,758) | T2DM with ASCVD (N = 22,680) | T2DM with HF (N = 3,027) | T2DM with ASCVD & HF (N = 8,051) |
|---|---|---|---|---|---|---|---|
| **All visits** | **Total visits** | 258557 | 119584 | 138973 | 82829 | 14106 | 42038 |
| | **Total visits per patient (Median [IQR])** | 0 (0-2) | 0 (0-2) | 1 (0-5) | 1 (0-4) | 1 (0-5) | 2 (0-6) |
| | **Annualized visits per patient (Median [IQR])** | 0.12 (0.0-0.56) | 0.0 (0.0-0.34) | 0.45 (0.0-1.19) | 0.36 (0.0-0.96) | 0.5 (0.0-1.31) | 0.84 (0.15-1.97) |
| **Inpatient Visits** | **Total visits** | 31940 | 12390 | 19550 | 10076 | 2114 | 7360 |
| | **Total visits per patient (Median [IQR])** | 0 (0-0) | 0 (0-0) | 0 (0-1) | 0 (0-0) | 0 (0-1) | 0 (0-1) |
| | **Annualized visits per patient (Median [IQR])** | 0.0 (0.0-0.0) | 0.0 (0.0-0.0) | 0.0 (0.0-0.14) | 0.0 (0.0-0.1) | 0.0 (0.0-0.17) | 0.0 (0.0-0.38) |
| **ED Visits** | **Total visits** | 16977 | 10370 | 6607 | 4205 | 663 | 1739 |
| | **Total visits per patient (Median [IQR])** | 0 (0-0) | 0 (0-0) | 0 (0-0) | 0 (0-0) | 0 (0-0) | 0 (0-0) |
| | **Annualized visits per patient (Median [IQR])** | 0.0 (0.0-0.0) | 0.0 (0.0-0.0) | 0.0 (0.0-0.0) | 0.0 (0.0-0.0) | 0.0 (0.0-0.0) | 0.0 (0.0-0.0) |
| **Outpatient Visits** | **Total visits** | 209640 | 96824 | 112816 | 68548 | 11329 | 32939 |
| | **Total visits per patient (Median [IQR])** | 0 (0-2) | 0 (0-1) | 1 (0-4) | 1 (0-3) | 1 (0-4) | 1 (0-4) |
| | **Annualized visits per patient (Median [IQR])** | 0.0 (0.0-0.42) | 0.0 (0.0-0.25) | 0.29 (0.0-0.89) | 0.25 (0.0-0.76) | 0.28 (0.0-0.94) | 0.48 (0.0-1.37) |

**Table S8. Financial burden associated with healthcare resource utilization due to cardiovascular causes across study cohorts.** Abbreviations: ASCVD, atherosclerotic cardiovascular disease; HF, heart failure; T2DM, type-2 diabetes mellitus.

| Type of Visit | Metric | Overall Population (N = 106,881) | T2DM w/o ASCVD & HF (N = 73,123) | T2DM with ASCVD or HF (N = 33,758) | T2DM with ASCVD (N = 22,680) | T2DM with HF (N = 3,027) | T2DM with ASCVD & HF (N = 8,051) |
|---|---|---|---|---|---|---|---|
| **All visits** | **Overall charges** | $795.1M | $323.9M | $471.3M | $245.7M | $57.3M | $168.2M |
| | **Total charges per patient (Median [IQR])** | $0 (0-224) | $0 (0-112) | $82 (0-7023) | $82 (0-672) | $112 (0-11558) | $123 (0-15923) |
| | **Annualized charges per patient (Median [IQR])** | $6 (0-53) | $0 (0-28) | $31 (0-2048) | $24 (0-789) | $42 (0-2703) | $94 (7-6204) |
| **Inpatient Visits** | **Overall charges** | $780.6M | $316.5M | $464.2M | $241.5M | $56.5M | $166.2M |
| | **Total charges per patient (Median [IQR])** | $0 (0-0) | $0 (0-0) | $0 (0-6952) | $0 (0-0) | $0 (0-10759) | $0 (0-15841) |
| | **Annualized charges per patient (Median [IQR])** | $0 (0-0) | $0 (0-0) | $0 (0-1975) | $0 (0-650) | $0 (0-2641) | $0 (0-6155) |
| **ED Visits** | **Overall charges** | $1.4M | $0.9M | $0.5M | $0.3M | $0.1M | $0.1M |
| | **Total charges per patient (Median [IQR])** | $0 (0-0) | $0 (0-0) | $0 (0-0) | $0 (0-0) | $0 (0-0) | $0 (0-0) |
| | **Annualized charges per patient (Median [IQR])** | $0 (0-0) | $0 (0-0) | $0 (0-0) | $0 (0-0) | $0 (0-0) | $0 (0-0) |
| **Outpatient Visits** | **Overall charges** | $13.1M | $6.5M | $6.6M | $4.0M | $0.7M | $1.9M |
| | **Total charges per patient (Median [IQR])** | $0 (0-112) | $0 (0-112) | $41 (0-205) | $41 (0-205) | $41 (0-224) | $41 (0-224) |
| | **Annualized charges per patient (Median [IQR])** | $0 (0-25) | $0 (0-16) | $15 (0-48) | $13 (0-41) | $16 (0-55) | $25 (0-74) |

**Table S9. Proportion of people at risk for financial hardship and catastrophic healthcare expenses.** Abbreviations: ASCVD, atherosclerotic cardiovascular disease; HF, heart failure; T2DM, type-2 diabetes mellitus.

| Population | Metric | Overall Population | T2DM w/o ASCVD & HF | T2DM with ASCVD or HF | T2DM with ASCVD | T2DM with HF | T2DM with ASCVD & HF |
|---|---|---|---|---|---|---|---|
| Study Population | **Total Number** | 106,881 | 73,123 | 33,758 | 22,680 | 3,027 | 8,051 |
| | **Patients facing financial hardship** | 12466 (11.66%) | 6262 (8.56%) | 6204 (18.38%) | 3182 (14.03%) | 713 (23.55%) | 2309 (28.68%) |
| | **Patients facing catastrophic healthcare expenses** | 6352 (5.94%) | 2980 (4.08%) | 3372 (9.99%) | 1581 (6.97%) | 407 (13.45%) | 1384 (17.19%) |
| Patients who died during follow-up | **Total Number** | 9587 | 4020 | 5567 | 2896 | 593 | 2078 |
| | **Patients facing financial hardship in the final year of life** | 5508 (57.45%) | 2250 (55.97%) | 3258 (58.52%) | 1626 (56.15%) | 350 (59.02%) | 1282 (61.69%) |
| | **Patients facing catastrophic healthcare expenses in the final year of life** | 3788 (39.51%) | 1579 (39.28%) | 2209 (39.68%) | 1080 (37.29%) | 247 (41.65%) | 882 (42.44%) |

**Table S10. Healthcare utilization and financial burden among patients who died during the study period.** Abbreviations: ASCVD, atherosclerotic cardiovascular disease; HF, heart failure; T2DM, type-2 diabetes mellitus.

| Type of Visit | Time Duration | Metric | Overall Population (N = 9587) | T2DM w/o ASCVD & HF (N = 4020) | T2DM with ASCVD or HF (N = 5567) | T2DM with ASCVD only (N = 2896) | T2DM with HF only (N = 593) | T2DM with ASCVD & HF (N = 2078) |
|---|---|---|---|---|---|---|---|---|
| All visits | Study period | Overall visits | 268183 | 124872 | 143311 | 75825 | 16859 | 50627 |
| | | Total visits per patient (Median [IQR]) | 17 (7-36) | 19 (8-39) | 15 (6-34) | 16 (6-35) | 17 (6-40) | 14 (6-31) |
| | | Overall charges | $888.8M | $369.5M | $519.3M | $243.8M | $68.4M | $207.2M |
| | | Total charges per patient (Median [IQR]) | $14336 (4654-34204) | $11684 (3562-28375) | $16372 (5601-38822) | $13188 (4304-30331) | $18928 (5731-45468) | $21704 (8383-49416) |
| | Final year of life | Overall visits | 90702 | 41877 | 48825 | 25196 | 5478 | 18151 |
| | | Total visits per patient (Median [IQR]) | 5 (2-12) | 6 (2-13) | 5 (2-11) | 5 (2-11) | 6 (2-11) | 6 (2-12) |
| | | Overall charges | $477.9M | $204.5M | $273.5M | $128.2M | $36.4M | $108.8M |
| | | Total charges per patient (Median [IQR]) | $25976 (1348-59104) | $25072 (672-60483) | $26765 (6815-58246) | $24380 (728-55563) | $29089 (7624-63441) | $29807 (9778-62464) |
| Inpatient Visits | Study period | Overall visits | 37873 | 14675 | 23198 | 10872 | 2706 | 9620 |
| | | Total visits per patient (Median [IQR]) | 3 (1-5) | 2 (1-5) | 3 (1-6) | 2 (1-5) | 3 (1-6) | 3 (1-6) |
| | | Overall charges | $875.4M | $362.5M | $512.9M | $240.4M | $67.5M | $205.0M |
| | | Total charges per patient (Median [IQR]) | $13984 (4432-33688) | $11346 (3257-27908) | $16094 (5357-38420) | $12920 (4121-30042) | $18610 (5513-45301) | $21548 (8127-48907) |
| | Final year of life | Overall visits | 19001 | 7617 | 11384 | 5364 | 1280 | 4740 |
| | | Total visits per patient (Median [IQR]) | 2 (1-3) | 2 (1-3) | 2 (1-4) | 2 (1-3) | 2 (1-4) | 2 (1-4) |
| | | Overall charges | $473.9M | $202.4M | $271.6M | $127.2M | $36.2M | $108.2M |
| | | Total charges per patient (Median [IQR]) | $39084 (19910-76411) | $39904 (19910-77477) | $38765 (20070-73530) | $37589 (18973-70246) | $40809 (20298-77477) | $39907 (21898-76674) |

| | | | | | | | | |
|---|---|---|---|---|---|---|---|---|
| **ED visits** | **Study period** | **Overall visits** | 23691 | 10751 | 12940 | 6644 | 1550 | 4746 |
| | | **Total visits per patient (Median [IQR])** | 1 (0-3) | 1 (0-3) | 1 (0-3) | 1 (0-3) | 1 (0-3) | 1 (0-3) |
| | | **Overall charges** | $1.6M | $0.8M | $0.8M | $0.4M | $0.1M | $0.3M |
| | | **Total charges per patient (Median [IQR])** | $13 (0-42) | $12 (0-42) | $13 (0-41) | $11 (0-37) | $15 (0-53) | $15 (0-46) |
| | **Final year of life** | **Overall visits** | 7212 | 3118 | 4094 | 2041 | 457 | 1596 |
| | | **Total visits per patient (Median [IQR])** | 1 (1-2) | 1 (1-2) | 1 (1-2) | 1 (1-2) | 1 (1-2) | 1 (1-2) |
| | | **Overall charges** | $0.4M | $0.2M | $0.2M | $0.1M | $0.0M | $0.1M |
| | | **Total charges per patient (Median [IQR])** | $82 (41-112) | $82 (41-123) | $41 (41-112) | $41 (41-112) | $82 (41-123) | $41 (41-112) |
| **Outpatient Visits** | **Study period** | **Overall visits** | 206619 | 99446 | 107173 | 58309 | 12603 | 36261 |
| | | **Total visits per patient (Median [IQR])** | 11 (3-27) | 13 (5-31) | 9 (3-25) | 10 (3-26) | 9 (3-28) | 8 (2-22) |
| | | **Overall charges** | $11.8M | $6.2M | $5.6M | $3.0M | $0.7M | $1.9M |
| | | **Total charges per patient (Median [IQR])** | $158 (61-374) | $177 (72-417) | $145 (54-345) | $141 (53-325) | $160 (51-419) | $149 (55-354) |
| | **Final year of life** | **Overall visits** | 64489 | 31142 | 33347 | 17791 | 3741 | 11815 |
| | | **Total visits per patient (Median [IQR])** | 4 (2-11) | 5 (2-12) | 4 (2-10) | 4 (2-10) | 4 (2-10) | 4 (2-9) |
| | | **Overall charges** | $3.6M | $1.9M | $1.7M | $0.9M | $0.2M | $0.6M |
| | | **Total charges per patient (Median [IQR])** | $205 (82-560) | $246 (112-672) | $205 (82-451) | $205 (82-451) | $205 (82-574) | $205 (82-451) |
